# Precision Transfusion Management: Rh Phenotype Compatibility and Antibody Surveillance in Southern China

**DOI:** 10.64898/2026.08.05.26359788

**Authors:** Xiao-qing Huang, Liang-Xing Li, Zhi-Yong Yang, Xing-Xing Long, Cai-Yun Lai

## Abstract

**Objective:** To analyze Rh antigen (C, c, D, E, e) distribution and alloimmunization patterns in Hengyang, China, and establish precision transfusion strategies.

**Methods:** Rh phenotyping and antibody screening were performed on 3,635 hospitalized patients and 5,326 donors using serological cards. A transfusion management system tracked historical antibodies and flagged discrepancies.

**Results:** Antigen frequencies in patients were D (99.56%) > e (94.69%) > C (91.64%) > c (48.06%) > E (38.79%). Dominant Rh(D)-positive phenotypes included CCDee (51.31%) and CcDEe (30.01%). After implementing Rh-matched transfusions (March–October 2023), antibody screening positivity decreased from 1.14% (2022) to 0.97% (p < 0.05). Rh system antibodies accounted for 46.01% (127/276) of positives in 2023, down from 55.21% in 2022 (p < 0.05), and were the leading cause of crossmatch incompatibility.

**Conclusion:** Rh antigen-matched transfusions significantly reduce alloimmunization risks. Continuous antibody tracking via transfusion management systems enhances transfusion safety.

## 1. Introduction

The International Society of Blood Transfusion (ISBT) has identified 43 blood group systems and 349 red blood cell antigens [1]. A blood group system is defined as a genetically discrete system comprising one or more antigens determined by a single gene or a complex of closely linked homologous genes [2]. ABO and Rh systems have long been regarded as the most important systems in blood transfusion medicine and transplantation [3–7]. Over 50 Rh antigens have been identified [8–10], with five exhibiting the strongest clinical relevance, ranked by antigenicity from strongest to weakest: D > E > C > c > e [11–12]. Clinical Transfusion Technical Specifications currently require ABO and RhD identical transfusions in healthcare institutions. However, no explicit guidelines exist for Rh C, c, E, and e antigen matching, which increases the risk of unexpected antibody formation in recipients. Unexpected antibodies, predominantly Rh system antibodies (accounting for over 46% [13–14]), are frequently induced by allogeneic transfusions or pregnancy and may lead to severe adverse events such as hemolytic transfusion reactions [15–17] and hemolytic disease of the fetus and newborn (HDFN) [18–20].

In this study, we analyzed the Rh phenotype distribution among 3,635 hospitalized patients and 5,326 blood donors from Hengyang, China, by integrating Rh phenotype serological testing, antibody screening, and real-time monitoring of Rh antigen-compatible transfusions through a blood transfusion management system. The objectives were to (1) establish regional Rh phenotype distribution frequencies, (2) quantify the incidence of Rh-related antibodies in patients requiring recurrent transfusions, and (3) evaluate the effectiveness of Rh antigen-compatible transfusions in reducing the production of unexpected antibodies. Our findings demonstrate that implementing Rh phenotype testing and precision Rh-matched transfusions significantly reduces the antibody screening positivity rate, serving as a critical strategy for preventing Rh alloimmunization in patients undergoing repeated transfusions. These results underscore the necessity for clinical transfusion services to prioritize compatibility for the four additional Rh antigens (C, c, E, and e) in addition to RhD.

## 2. Materials and Methods

### 2.1 Study Population

This cross-sectional study was designed and executed in Hengyang. A total of 28,594 patients who received blood transfusions from March 2023 to October 2023 were included, comprising 12,938 males and 15,656 females. Among them, 3,635 patients received ABO and RhD/C/c/E/e antigen-compatible transfusions. As a control, 27,886 patients transfused between March 2022 and October 2022 were retrospectively analyzed (12,438 males and 15,448 females), who received ABO/RhD-matched transfusions with Rh C/c/E/e phenotypes unknown (Table 1). In addition, Rh phenotyping was also performed on 5,326 blood donor samples. This retrospective study utilized existing clinical data and biological samples, with an informed consent approved by our Hospital (Approval No.: 2023LL0512001).

**Table 1.** Demographic features of the participants.

| Year | Male (% (Count)) | Female (% (Count)) | Mean Age |
| --- | --- | --- | --- |
| 2022 | 44.6 (12,438) | 55.4 (15,448) | 57.5 |
| 2023 | 45.2 (12,938) | 54.8 (15,656) | 55.2 |

### 2.2. Research Methods

#### 2.2.1. Rh Phenotyping

Rh antigens (C, c, D, E, e) were preliminary screening and confirmation using microcolumn gel Rh phenotyping cards (Shanghai Runpu Biotechnology Co., Ltd., China; Bio-Rad Laboratories, USA) method prior to the first transfusion and recorded in the Clinical Blood Transfusion Quality Management System (Hunan Innovation Technology Co., Ltd., China) as a reference. For subsequent transfusions, Rh antigens were re-evaluated and compared with initial results. Abnormal results (e.g., trailing or mixed-field reactions) were confirmed using capillary high-speed separation techniques (Wuhan beisuo medical equipment co., ltd, China).

#### 2.2.2. Antibody Screening and Identification

Unexpected antibodies were screened using the IH-1000 automated analyzer (Bio-Rad Laboratories, USA) with Antibody screening cells (3-cell panel, Shanghai Blood Biopharmaceutical Co., Ltd., China) and low-ionic-strength antihuman globulin cards (Bio-Rad Laboratories, USA). Antibody-positive samples were recorded in the Clinical Blood Transfusion Quality Management System. Antibody specificity was further identified using red blood cell antibody identification cells (10-cell panel, Shanghai Blood Biopharmaceutical Co., Ltd., China) via microcolumn gel antihuman globulin (Bio-Rad Laboratories, USA) and/or saline methods. For weak or ambiguous antibodies, enhancement techniques (e.g., Liss-IAT) were applied, and antibody distribution was statistically analyzed.

#### 2.2.3. Continuous Tracking of Blood Group Antibodies and Antigens

The Clinical Blood Transfusion Quality Management System documented patients’ ABO, RhD, Rh phenotypes, and specific antibodies. When discrepancies between current and historical results were detected, the system automatically alerted clinicians to the historical antigen and antibody profiles and recommended isotype-compatible blood transfusions tailored to the identified antibodies.

#### 2.2.4. Statistical Analysis

Data were analyzed using IBM SPSS Statistics for Windows, Version 21.0. Armonk, NY: IBM Corp. Chi-square tests were applied, with *p* < 0.05 considered statistically significant.

## 3. Results

### 3.1. Distribution of Major Rh Antigens

Rh antigen typing (C, c, D, E, e) was performed in 3,635 hospitalized patients and 5,326 blood donors. The antigen frequency trend in patients was D (99.56%) > e (94.69%) > C (91.64%) > c (48.06%) > E (38.79%), while in donors, the trend was D (99.76%) > e (94.33%) > C (91.53%) > c (49.27%) > E (40.27%). No statistically significant differences were observed between patients and donors (*p* > 0.05).

### 3.2. Rh Phenotype Distribution

Among 3,635 patients, 3,619 were Rh(D)-positive, with nine phenotypes detected: CCDee (51.31%) > CcDEe (30.01%) > CcDee (9.37%) > ccDEE (5.00%) > ccDEe (2.79%) > CCDEe (0.80%) > ccDee (0.39%) > CcDEE (0.28%) > CCDEE (0.05%). The 16 Rh(D)-negative patients exhibited phenotypes: ccee (8 cases) > Ccee (7 cases) > CcEe (1 case). Similarly, 5,313 Rh(D)-positive donors showed comparable phenotype frequencies: CCDee (49.99%) > CcDEe (31.07%) > CcDee (9.41%) > ccDEE (5.50%) > ccDEe (2.75%) > CCDEe (0.85%) > ccDee (0.24%) > CcDEE (0.17%) > CCDEE (0.02%). The phenotypic distribution between patients and donors was consistent (*p* > 0.05).

### 3.3. Antibody Screening Results

From March to October 2022, 317 out of 27,886 patients (1.14%) tested positive for antibodies. In 2023, 276 out of 28,594 patients (0.97%) were antibody-positive, showing a statistically significant decrease (*p* < 0.05).

### 3.4. Specific Antibody Identification

Among 276 antibody-positive patients in 2023, 127 (46.01%) had Rh system antibodies. Anti-E was predominant (34.78%, 96/276), followed by anti-c/cE (5.07%, 14/276) and anti-D (3.62%, 10/276). Notably, the Anti-D detection rate in 2023 (3.62%) showed an upward trend compared to the 2022 (3.47%) and previous reports [21–22]. This increase may be attributed to enhanced awareness of Anti-D antibodies among RhD-negative pregnant women, who underwent Anti-D titer monitoring during prenatal care. Medical records indicated that some Anti-D-positive pregnant women in 2023 received prophylactic anti-D immunoglobulin, resulting in the production of protective Anti-D antibodies. Other findings included MNSs-system antibodies (14.13%, 39/276), autoantibodies (14.13%, 39/276), and anti-drug antibodies (4.35%, 12/276). Rh-related antibody prevalence in 2023 (46.01%) was significantly lower than in 2022 (55.21%, *p* < 0.05) (Table 2).

**Table 2.** The distribution of specific antibodies among hospitalized patients.

| Specific Antibody |  | 2023 (n = 276) | 2022 (n = 317) |
| --- | --- | --- | --- |
|  |  | (% (count)) | (% (count)) |
| Rh | Anti-E | 34.78 (96) | 40.06 (127) |
|  | Anti-c/cE | 5.07 (14) | 9.15 (29) |
|  | Anti-D | 3.62 (10) | 3.47 (11) |
|  | Anti-c/Ce | 2.54 (7) | 2.52 (8) |
| MNSs-system | Anti-M | 14.13 (39) | 16.72 (53) |
| Lewis-system | Anti-Lea | 2.54 (7) | 2.84 (9) |
| P-system | Anti-P1 | 1.81 (5) | 1.25 (4) |
| Kidd-system | Anti-Jka | 0.36 (1) | 0.32 (1) |
| Autoantibody |  | 14.13 (39) | 9.46 (30) |
| Anti-drugs antibody |  | 4.35 (12) | 3.16 (10) |
| Others |  | 16.67 (45) | 11.04 (35) |

### 3.5. Departmental Distribution of Antibody-Positive Patients

Rh system antibodies (46.01%, 127/276) were predominantly observed in hematology, ICU, hepatobiliary surgery, and gastroenterology departments, primarily among patients requiring repeated transfusions (Table 3).

**Table 3.** Distribution of antibody screening positive patients across departments.

| Department | 2023 (n = 276) | Rh Antibody<br>(count (%)) | 2022 (n = 317) | Rh Antibody<br>(count (%)) |
| --- | --- | --- | --- | --- |
| Obstetrics | 14 | 11 (78.57) | 15 | 12 (80.00) |
| Gynecology | 16 | 9 (56.25) | 14 | 8 (57.14) |
| Hematology | 52 | 21 (40.38) | 70 | 48 (68.57) |
| Cardiology | 3 | 3 (100.00) | 1 | 1 (100.00) |
| Cardiovascular<br>Surgery | 14 | 7 (50.00) | 15 | 8 (53.33) |
| Emergency<br>Department | 7 | 5 (71.43) | 12 | 10 (83.33) |
| Infectious<br>Diseases | 5 | 3 (60.00) | 0 | 0 (0) |
| Urology | 23 | 10 (43.48) | 24 | 14 (58.33) |
| Gastroenterology | 19 | 12 (63.16) | 30 | 20 (66.67) |
| Neurology | 3 | 1 (33.33) | 5 | 2 (40.00) |
| Neurosurgery | 3 | 3 (100.00) | 6 | 3 (50.00) |
| Hepatobiliary<br>Surgery | 21 | 10 (47.62) | 21 | 8 (38.10) |
| Nephrology | 12 | 9 (75.00) | 2 | 2 (100.00) |
| Gastrointestinal | 10 | 3 (30.00) | 21 | 9 (42.86) |
| Surgery |  |  |  |  |
| Spinal Surgery | 14 | 6 (42.86) | 9 | 4 (44.44) |
| ICU | 23 | 10 (43.48) | 15 | 7 (46.67) |
| Orthopedics | 19 | 3 (15.79) | 21 | 6 (28.57) |
| Rheumatology &<br>Immunology | 9 | 1 (11.11) | 4 | 1 (25.00) |
| Pediatrics | 7 | 0 (0) | 5 | 1 (20.00) |
| Breast &<br>Lymphoma Ward | 2 | 0 (0) | 10 | 6 (60.00) |
| EICU | 0 | 0 (0) | 7 | 3 (42.86) |
| Oncology | 0 | 0 (0) | 10 | 2 (20.00) |

### 3.6. Inconsistent Historical Antibody Alerts

The transfusion system flagged 25 cases (2023) with inconsistent historical antibody results, of which 60% (15/25) involved specific antibodies, predominantly Rh system (56%, 14/25), indicating declining antibody titers over time (Table 4).

**Table 4.** Distribution of specific antibodies with inconsistent historical antibody screening results.

| Specific Antibody |  | Patients (n = 25) |
| --- | --- | --- |
|  |  | (% (count)) |
| Rh system | Anti-E | 48 (12) |
|  | Anti-D | 4 (1) |
|  | Anti-C | 4 (1) |
| Lewis-system | Anti-Lea | 4 (1) |
|  | Autoantibody | 40 (10) |

### 3.7. Rh Phenotype Compatibility Probability

Based on the principles of phenotypic compatibility and phenotype frequencies, we calculated the Rh phenotype transfusion compatibility probabilities (Table 5). Phenotypic compatibility is defined as either: Complete identity of Rh phenotypes (D, C, c, E, e) between donor and recipient, or Recipient Rh phenotype containing all antigens present in the donor phenotype, even if not fully identical. These blood compatibility probabilities reflect the likelihood that the current blood inventory in transfusion departments can achieve compatible transfusions between donors and recipients, while also providing critical insights for optimizing blood inventory allocation strategies.

**Table 5.** Rh phenotype compatibility (%)

| Phenotype | Frequency | Compatibility |
| --- | --- | --- |
| CCee | 49.99 | 49.99 |
| CcEe | 31.07 | 100.00 |
| Ccee | 9.41 | 59.64 |
| ccEE | 5.50 | 5.50 |
| ccEe | 2.75 | 8.49 |
| CCEe | 0.85 | 50.86 |
| Ccee | 0.24 | 0.24 |
| CcEE | 0.17 | 5.69 |
| CCEE | 0.02 | 0.02 |

## 4. Discussion

Most Rh antibodies are IgG1 subclass, with some IgG2/IgG3 [23], implicated in delayed hemolytic transfusion reactions and HDFN [24]. Among 3,635 hospitalized patients and 5,326 donors, nine Rh(D)-positive phenotypes were identified, with CCDee (51.31%) and CcDEe (30.01%) predominating (collectively >81%). Antigen frequencies in patients followed D (99.56%) > e (94.69%) > C (91.64%) > c (48.06%) > E (38.79%), aligning with donors (*p* > 0.05) and prior reports [25]. These findings highlight elevated risks of anti-E and anti-c formation without Rh-compatible transfusions. Notably, anti-E, the second most common cause of HDFN in China [26], underscores the clinical urgency.

Unexpected antibodies from Rh, MNS, Lewis, Duffy, Kidd, and P systems remain key contributors to crossmatch incompatibility [27–31]. In this study, Rh system antibodies accounted for 46.01% of cases. Following Rh phenotyping and antigen-compatible transfusions in 2023, antibody screening positivity decreased from 1.14% (2022) to 0.97% (*p* < 0.05), with Rh-specific antibody prevalence declining from 55.21% to 46.01%. These trends reinforce the importance of Rh phenotyping and longitudinal antibody tracking.

According to the requirements of relevant testing standards, Rh blood group system antigen identification should be carried out before blood transfusion [32]. Currently, Rh-matched transfusions have not been universally adopted across Chinese healthcare institutions. To address this gap, our transfusion department established an efficient blood transfusion information system. During a patient’s first transfusion, their ABO and RhD blood types, Rh antigen phenotypes, and irregular antibodies are recorded in the system. Transfusion and pregnancy-related alloimmunization are the primary stimuli for the development of irregular antibodies. The probability of generating irregular antibodies in recipients is approximately 0.4% per 10 units of red blood cells transfused [33]. A hallmark of many irregular antibodies is the gradual decline or eventual disappearance of antibody titers over time post-induction [34]. Leveraging the transfusion management system, this study implemented an antibody registry to meticulously track the lifecycle of antibodies - from emergence to decline and resolution. Consequently, when antibody titers fall below detectable thresholds, the system alerts clinicians to historical antibody types and Rh antigen phenotypes in recurrent transfusion recipients. This approach enhances the timeliness of complex antibody detection, mitigates Rh system alloimmunization risks, and enables precision transfusion strategies tailored to individual patient profiles.

Currently, some medical institutions in China are gradually implementing Rh-compatible transfusions, which require close collaboration with local blood banks. However, only a limited number of blood banks provide Rh phenotyping results for red blood cells, necessitating simultaneous Rh antigen typing for both patients and donors in transfusion departments. This imposes significant testing workload challenges. Given the nationwide shortage of blood supply, universal protocols can be adopted to ensure transfusion safety when conducting Rh-compatible transfusions. Based on phenotype compatibility principles, this study calculated the compatibility probabilities for different Rh antigen phenotypes.

Phenotypes such as CCee, CcEe, Ccee, and CCEe demonstrated compatibility probabilities exceeding 50%, indicating that matching blood units are readily available in most transfusion department inventories. In contrast, low-frequency phenotypes (e.g., ccEe, ccEE, CcEE) showed compatibility probabilities of approximately 5%, requiring access to medium- or large-scale blood inventories. Rare phenotypes (e.g., ccee, CCDEE) exhibited compatibility probabilities below 1%, necessitating advance coordination with regional blood centers to source units from rare blood type registries. These compatibility probabilities provide critical guidance for optimizing blood inventory capacity in transfusion departments, thereby enhancing the efficiency of emergency transfusion protocols and improving clinical outcomes. A limitation is the reliance on serological methods; future work will integrate genetic analyses (e.g., RhD variants, adsorption-elution genotypes) to deepen biological insights.

## 5. Conclusions

This study provides regional Rh phenotype distribution data and validates Rh-compatible transfusion protocols. Implementing Rh phenotyping and antigen-compatible transfusions reduces antibody prevalence, offering a robust strategy against alloimmunization. Medical institutions should pay greater attention to Rh antigen (C, c, E, and e) compatibility in clinical blood transfusion practice.

## Data Availability

All data produced in the present study are available upon reasonable request to the authors

## Abbreviations

ISBT: International Society of Blood Transfusion
HDFN: hemolytic disease of the fetus and newborn

